# Assessing the implementation of user-centred design standards on assistive technology for persons with visual impairments: A systematic review

**DOI:** 10.1101/2023.03.10.23287090

**Authors:** Luisa María Ortiz-Escobar, Mario Andres Chavarria, Klaus Schönenberger, Samia Hurst-Majno, Michael Ashley Stein, Anthony Mugeere, Minerva Rivas-Velarde

## Abstract

Despite scientific and technological advances in the field of assistive technology (AT) for people with visual impairment (VI), technological designs are frequently based on a poor understanding of the physical and social context of use, resulting in devices that are less than optimal for their intended beneficiaries. To resolve this situation, user-centred approaches in the development process of AT have been widely adopted in recent years. However, there is a lack of systematization on the application of this approach. This systematic review registered in PROSPERO (CRD42022307466), assesses the application of the ISO 9241-210 human-centred design principles in allegedly “user-centred designed” AT developments for persons with VI (see S1 PROSPERO Protocol). The results point to a wide variation of the depth of understanding of user needs, a poor characterization of the application of the User Centred Design (UCD) approach in the initial design phases or in the early prototyping, and a vague description of user feedback and device iteration. Among the principles set out in ISO 9241-210, the application of 5.6: “the design team includes multidisciplinary skills and perspectives” is the one for which the least evidence is found. The results show there is not enough evidence to fully assess the impact of UCD in 1. promoting innovation regarding AT products and practices, and 2. Judging if AT produced following such standards is leading to better user access, wellbeing outcomes and satisfaction. To address this gap it is necessary to, first, generate better implementation of UCD in AT development and second, to strengthen evidence regarding the implementation and outcomes of using UCD for AT. To better engage with the realities of persons with VI, we propose capacity building across development teams regarding UCD, its principles and components; better planning for UCD implementation; and cross-fertilization across engineering disciplines and social and clinical science.

## Introduction

User-Centred Design (UCD) has gained a stronger presence in Assistive Technology (AT) development over the last decade [1]. This approach promotes the involvement of end users in all stages of the design process, elicitation and understanding of their needs, and characterization of social contexts as the basis for an iterative design process [2–3]. Therefore, UCD adoption is believed to lead to better products [4]. However, there is limited evidence regarding the implementation of these approaches or if their results are having the intended impact across their target populations, particularly regarding AT [5–6]. This study aims to assess the application of ISO 9241-210 human-centred design principles in the allegedly “user-centred designed” assistive technology developments for persons with Visual Impairments (VI).

The Global Report on Assistive Technology (GREAT) states that children and adults with disabilities lack access to AT, particularly in low-and-middle-income Countries (LMICs) where access was reported to be as low as 3% [7]. The current lack of access to AT reflects not only an economic gap but a severe malfunction of social provision and coverage schemes as well as in AT design and development [8]. Nevertheless, UCD and international standards’ adoption can help to alleviate these shortfalls by guiding the development of better and more efficient AT solutions responding to the users’ priorities. Disability is very diverse and persons with different impairments, namely sensorial, physical or cognitive or multiples, benefit from different technological solutions; we need to learn more about similarities, as well as differences. Therefore, in this paper, the focus is on AT for persons with VI. Worldwide, there are approximately 39 million people with severe VI or blindness [9]. Although not all-disabling loss of sight can be addressed by AT, for persons who are blind (visual acuity worse than 3/60) some tools such as walking canes, screen readers, or braille embossers, amongst others, are of great help. The Global report on disability calls for action and standard setting in a variety of AT related fields, particularly regarding access [7]. Thus, investigating how internationally adopted standards are implemented for technology design is relevant to close the AT gap. The upcoming section explores relevant international standards for the production of AT.

### AT and International Standards

International standards play a key role in the development, production and distribution of technology [10]. The existence of clear, accessible, and commonly accepted International Standards is vital for the manufacture of products that can be globally implemented and commercialized. Standardization enhances product quality, safety, and reliability, it also allows for higher interoperability and compatibility in different contexts and reduces maintenance complexity and costs [11]. There are different studies on the positive effect of international standardization on trade, industry and management [10, 12], including evidence of the reduction of barriers to the export and acceptance of products between different global regions, including products exported from LMICs to high income Countries [13]. Given the current AT access gap and lack of evidence on how available technology responds to the needs of persons with VI in LMIC it is relevant to look at how and whether the adoption of these standards can lead to better and more efficient AT. Furthermore, infrastructure for AT production and a well-defined value chain have an impact on AT access, nonetheless, this is outside the scope of this paper.

### The Standard of User-Centred Design

UCD is recognized by the International Organization for Standardization (ISO) in their standard ISO 9241-210, where it is described as an “approach to system design and development that aims to make interactive systems more usable by focusing on system use and applying human factors/ergonomics and usability knowledge and techniques” [3]. The standard presents a framework giving examples of activities that can be developed when adopting the approach. Furthermore, it clarifies that UCD is complementary to existing design methodologies, for example regarding usability [14] and Measurement of quality in use ISO/IEC 25022, amongst others. UCD is guided by the following 6 principles: (I) the design is based upon an explicit understanding of users, tasks and environments; (II) users are involved throughout design and development; (III) the design is driven and refined by user-centred evaluation; (IV) the process is iterative; (V) the design addresses the whole user experience; and (VI) the design team includes multidisciplinary skills and perspectives.

There is very narrow empirical evidence on the impact of standards on innovation, particularly regarding AT [15]. However, forthcoming empirical literature shows a positive influence of standards on the diffusion of technical knowledge and their contribution to macroeconomic growth. For example, a set of studies performed within different countries showed that the contribution of standards to the growth rate in each of the evaluated countries was equivalent to “0.9% in Germany, 0.8% in France and Australia, 0.3% in the UK and 0.2% in Canada” [12]. Another set of studies, performed by the ISO in several companies from different sectors in ten countries, showed an overall increase between 0.5% to 4% in the companies’ annual sales revenues provided by the implementation of international standards [13,16 -17].

The adoption of a user-centred design approach in the development process of AT has increased in recent years. This systematic review assesses the application of the ISO 9241-210 Human-centred design principles in the “user-centred designed” AT developments for persons with VI. The goal is to better understand how systematically the approach has been applied in the design and development of AT.

## Method

The present review followed the Prisma guidelines for systematic reviews seeking to answer the next question (S2 PRISMA Checklist) [18]

> *Do user-centred designed assistive technology developments for persons with visual impairments apply the human-centred design principles of the ISO 9241-210?*

The multidisciplinary databases Science Direct, Scopus, PubMed Central and Web of Science, were defined as primary sources. The electronic searches were performed in January 2022, and updated on June 2022. The keywords visual impairments (blindness and low vision), user-centred design, and assistive technology were used as search terms. The search period was established between January 2012 and March 2022. Considering that standards take time to become known and applied, a gap of two years was left between the publication of ISO 9241-210 (2010) and the start date of the search (2012). In any case, the application of the previous standard (ISO 13407) was considered during the revision. The complete Web of Science search strategy, was adapted for the other databases:

> Search string: ((“visual+ impair+” OR “visual+ disab+” OR blind OR “low vision”) AND (“user-centred design” OR “human-centred design” OR “ISO 9241-210” NOT “universal design”) AND (assistive technology))

### Eligibility Criteria

#### Inclusion criteria

- Topic of study: papers are describing the design and/or development process of user centred designed assistive technology for visually impaired persons..
- Type of scientific material to analyse: Peer-reviewed journals. Any type of research design: experimental, descriptive, or analytic research design. Except for letters and editorials. It will include systematic reviews and meta-analyses.
- Studies available in English / Spanish/ Portuguese / French
- Full text available
- Full conference papers

#### Exclusion criteria

- Articles that are not exclusively addressed to persons with VI.
- Articles describing assistive technology design or developments addressed for persons with VI without any consideration to the UCD approach.

### Study Selection

All search results were imported into an EndNote database. Duplicates were removed. Abstracts and titles that were noticeably unrelated to the review topic were dismissed. Two researchers independently screened the titles and abstracts against the eligibility criteria and selected those that met the inclusion criteria. Full-text reports were retrieved and again assessed for final eligibility. Reasons for excluding full-text reports were documented. The selected studies were analysed with a standardised data extraction form. Disagreements during the selection process were discussed in a consensus meeting with a third reviewer who helped to solve the discrepancies.

### Data Extraction

The following data was extracted from the selected studies:

- Data about the publication (authors, title of the article and the journal), aims, methods, design approaches (usability testing, workshops, interviews, focus groups, think-aloud, observation, including others.), frameworks, and studies design.
- Data about participants: sample size, socio-demographic characteristics, inclusion and exclusion criteria, type of impairments (low vision or blindness).
- Setting: country.
- And the application of ISO 9241-210’s principles in assistive technology developments.

### Data Quality Assessment

The quality assessment tool used in this review was drawn from Appendix D of Hawker et al. [19]. It was applied independently by two researchers to perform a risk of bias assessment, and disagreements were resolved through discussion with a third researcher. The overall quality grades definitions used by Lorenc et al. were adopted [20]. The tool was adapted to include the assessment of reviews. Only for this type of studies the scores were adjusted as follows: high quality (A), 27-32 points; medium quality (B), 22-26 points; low quality (C), 8-21 points (see S3).

### Data Analysis

All search results and their respective reasons for inclusion or exclusion were documented through The PRISMA flowchart. The body of evidence was analysed qualitatively by the themes stated in the “data extraction” regarding the ISO 9241-210’s principles application and is presented through a descriptive overview. The quantitative data (publication data, data about the participants, setting, and principles application) was analysed and presented through descriptive statistics tools.

### Protocol and Registration

The protocol describing this systematic review methodology was previously registered in PROSPERO (CRD42022307466).

## Results

Of the 348 references found in databases, 37 were fully analysed, and 28 were included for systematic review (See Fig 1).

**Fig 1.**
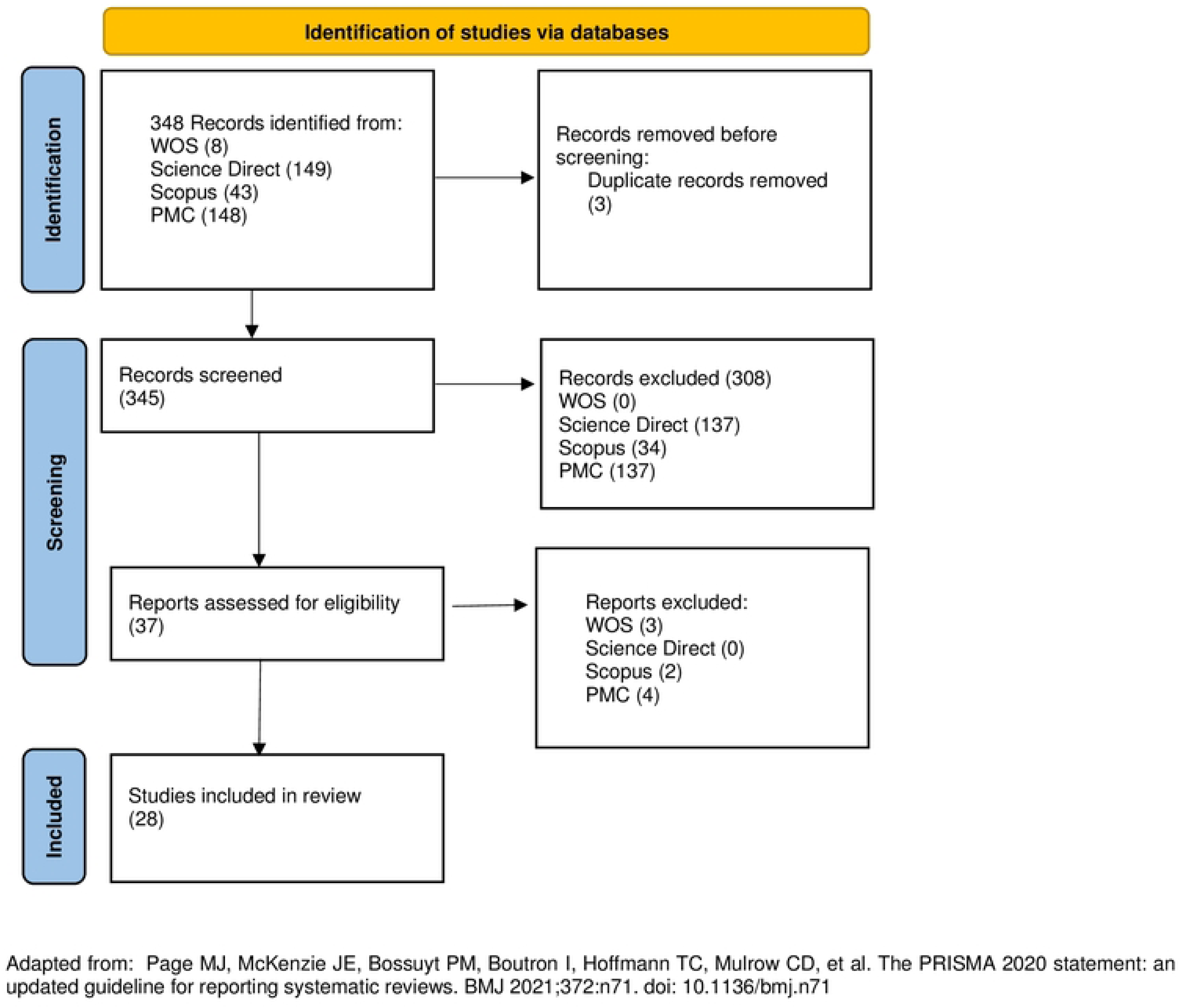
PRISMA flow diagram for eligible article identification

The aim of this systematic review is to examine and describe the application (or absence of application) of the ISO 9241-210’s principles in AT developments for persons with VI, based on searches in multidisciplinary databases. Here, we first provide a descriptive analysis of the papers outlining geographical representation and topics covered. Then, we present an in-depth analysis of findings pertaining to the development and adoption of the UCD principles.

### Themes covered in the literature and geo-representation

The results retrieved contributions from 16 countries. Regarding countries’ distribution by income category, it should be noted that 85.7% (24) of the papers included are from high-income countries, four from middle-income, and none from low-income countries (Table 1).

**Table 1.** Number of studies per country

| Country | N° of studies | Classification by income level |
| --- | --- | --- |
| Austria | 1 | High |
| Belgium | 1 | High |
| Brazil | 1 | Upper middle |
| Brazil-Chile | 1 | Upper middle - High |
| Chile | 1 | High |
| France | 2 | High |
| Germany | 1 | High |
| Greece | 1 | High |
| Italy | 2 | High |
| Malaysia | 1 | Upper middle |
| Portugal | 1 | High |
| Saudi Arabia | 2 | High |
| Spain | 2 | High |
| Taiwan | 1 | High |
| Thailand | 1 | Upper middle |
| UK | 2 | High |
| USA | 7 | High |

AT designs or developments covered the following areas (according to the ISO 9999:2016) [22]: activities and participation relating to personal mobility and transportation (ten papers); communication and information management (five papers); education and training in skills (six papers); work activities and participation in employment (one paper); and for assistive products for self-care activities and participation in self-care (two papers). Four of the 28 articles were literature reviews, three are oriented towards ATs for activities related to personal mobility and transportation, while the last one describes the usability of mobile applications for VIs to support different types of activities.

Most of the articles (21/28) explicitly declare using/implementing the UCD framework of reference. However, the implemented framework, namely the ISO 9241-210 (or its previous version ISO 13407) was referenced in only five of the reviewed articles [21, 23–26]. Excluding the aforementioned studies, only two papers [27–28], cited a reference, other than ISO 9241-210, for UCD, specifically (Cheverst et al. [29]) and (Nielsen [30]).

### Adoption of UCD principles

An analysis of the available evidence shows that papers documenting the development of AT for persons with VI tend to present a more detailed description of the state of the art in terms of the systems requirements than properly a characterization of the context of use or preferences and needs of the target user. Also, there is a strong emphasis on usability-oriented studies (64.29%). AT developments tend to engage users mostly at the end of the process to test if the product can be used. More than one third (35.71%) of the articles did exactly that. While 21.4% presented usability evaluation and results as part of the user-centred design process. Another 21.4% aimed to apply the user-centred design process in the early stages of the design or development process, looking at the feasibility of using the device and highlighting the compatibility and advantages of using participatory design methodologies with the UCD approach. The information extracted from the selected articles is presented below, grouped under each of the six principles of ISO 9241-210 to review their application (Table 2).

**Table 2.** Data extraction regarding the implementation of user-centred design standards on assistive technology for persons isual impairments.

| Authors | Article Title | Publication Year | Country | Aim | Sample | 5.2 The design is based upon an explicit understanding of users, tasks and environments | 5.3 Users are involved throughout design and development | 5.4 The design is driven and refined by user-centred evaluation | 5.5 The process is iterative | 5.6 The design addresses the whole user experience | 5.7 The design team includes multidisciplinary skills and perspectives | Data collection tools, techniques |
| --- | --- | --- | --- | --- | --- | --- | --- | --- | --- | --- | --- | --- |
| Conradie PD, De Marez L, Saldien J. [21] | Participation is Blind: Involving Low Vision Lead Users in Product Development | 2015 | Belgium | To present how the Lead User approach was applied, focusing on the tools and techniques we used. | Group 1 contained nine participants, while group 2 contained twelve. | Yes | Yes | Yes | Yes | Yes | Not described | Qualitative interviews with six (four females and two males) blind persons by phone, followed by two focus group discussions |
| Façanha AR, Araújo MC, Viana W, Sánchez J. [23] | Design and evaluation of mobile sensing technologies for identifying medicines by people with visual disabilities | 2019 | Brasil | To assist people with visual impairments to identify their medicine by using mobile sensing technologies. | "Expert" group (computing undergraduate students, an assistive technology researcher and an ophthalmologist); 2 blind adults; 10 VIP for the usability test | Yes | Yes | Yes | Iteration is described in the theoretical framework, not in the development process | Yes | This group had four computing undergraduate students, an assistive technology researcher and an ophthalmologist. | usability evaluation instruments (SUS and SUBC), accompanied by a brief interview about the users' impressions |
| Ntakolia C, Dimas G, Lakovidis DK. [24] | User-centered system design for assisted navigation of visually impaired individuals in outdoor cultural environments | 2020 | Greece | To develop a human-computer interactive system addressed to guide VIs in outdoor cultural environments. | 51 visually impaired, 77 non visually impaired for the questionnaire. For the evaluation process of the prototype 10 blinded users. | Yes | Users are involved in design and development, not in evaluation | Yes, but usability test was made with blinded participants | Yes | Yes | Not described | Observations in the outdoor environment, interviews, questionnaires, prototype usability test |
| Mattheiss E, Regal G, Sellitsch D, Tscheligi M. [25] | User-centred design with visually impaired pupils: A case study of a game editor for orientation and mobility training | 2016 | Austria | This aim raises open research questions on how to design an accessible O&M training game editor. | 25 teenagers and young adults from several classes of a school for VI pupils in Vienna, Austria, with business and polytechnical focus. | Yes | Yes | Yes | Yes | Yes | The design team is not described. | Semi-structured expert interview Workshop about brainstorming methods, digital survey, Focus group, Behaviour observation and contextual, exploratory interview in game playing, Self-experience in O&M training |
| Sánchez J. [26] | Development of Navigation Skills Through Audio Haptic Videogaming in Learners Who are Blind | 2012 | Chile | The purpose of this work was to investigate whether the use of audio and haptic-based videogame has an impact on the development of O&M skills in school-age blind learners. | For the usability evaluation: 10 blind learners (ages from 9-15 years). None of these participants have any additional, associated disabilities other than VI. Cognitive impact test: 7 blind learners (10 and 15 years old). | Yes | Yes | Yes | Yes | Yes | The research team is not described, Authors described abundant team experience regarding interface design for blind children was also used in this design process. | Interaction data was collected from twenty independent onsite user evaluations, using observation together with a think-aloud protocol, field notes, semi-structured interview and a specialized checklist. |
| Nimmolrat A, Khuwuthyakom P, Wientong P, Thinnukool O. [27] | Pharmaceutical mobile application for visually-impaired people in Thailand: development and implementation | 2021) | Thailand | The aim is to analyse, design and implement a mobile pharmaceutical application called "Ru Tan Ya" in the Thai language, which enables users to manage their medication and can be applied to both iOS and Android systems. | Sixty volunteers from the active members of the Vision Disability Association in northern Thailand were recruited. The inclusion criteria and sampling methods are presented. | Yes | Yes | Yes | no, but suggestions are made for improvements | Yes | not described | Interviews for the Identification of users' problems and needs. Feedback and ranking based in a usability questionnaire. |
| Yeh F, Yang C. [28] | Assisting the visually impaired to deal with telephone interview jobs using information and commutation technology | 2014 | Taiwan | The goal of this work was to develop a new ICT assisted blind telephone interview (ICT-ABTI) system to increase the working performance of the visually impaired. | Seven visually impaired people participated in this study. All of the participants graduated from university. All of the participants had enough experience in using computers. The ethnic backgrounds are presented. Participant's ages ranged from 24-39 years of age. | Yes | Yes | Yes | Yes | No | Not described | Interviews, This study adopted an ABAB design (Richards, Taylor, Ramasamy, & Richards, 1999) to execute the tests, in which A represented the baseline phase (without ICT-ABTI system) and B represented the intervention phase (with ICT-ABTI system). |
| Façanha AR, Darin T, Viana W, Sánchez J. [31] | O&M Indoor Virtual Environments for People Who Are Blind: A Systematic Literature Review | 2020 | Brazil-Chile | To systematically examine the literature on O&M virtual environments designed to support indoor navigation to identify techniques for both developing and evaluating the usability and cognitive impact of these applications. | 51 papers describing O&M indoor virtual environments | N/A: Literature review |  |  |  |  | Not described | Regarding evaluation techniques, questionnaires, interviews, user observation, and performance logs are commonly used to evaluate usability in this context. Most papers do not report any strategies to evaluate the cognitive impact of O&M virtual environments on users' navigational and wayfinding skills. |
| Hakobyan L, Lumsden J, O'Sullivan D, Bartlett H. [32] | Mobile assistive technologies for the visually impaired | 2013 | UK | To review research and innovation within the field of mobile assistive technology for the visually impaired and, in so doing, highlight the need for successful collaboration between clinical expertise, computer science, and domain users to realize fully the potential benefits of such technologies. | 168 articles were reviewed in detail | N/A: literature review |  |  |  |  | Authors highlighted the need for collaboration between clinical expertise and the field of computer science, but the research team behind the review is not described | "UCD encourages the use of user-focused design tools and practices including interviews, focus groups, surveys, usability testing, and participatory design processes." |
| Mascetti S, Picinali L, Gerino A, Ahmetović D, Bernareggi C. [33] | Sonification of guidance data during roadcrossing for people with visual impairments or blindness | 2015 | Italy | Two original auditory guiding modes based on data sonification are presented and compared with a guiding mode based on speech messages | Three sets of empirical evaluations were conducted: quantitative evaluation with 11 blindfolded sighted test subjects, a qualitative evaluation with 12 blind test subjects and, finally, a quantitative and qualitative evaluation conducted with 3 test subjects with VIB. | No | Users involved in preliminary and usability tests | No | Yes | No | The design team is not described. | Informal evaluations, preliminary evaluations. Experimental tests, quantitative variables related with time responses to the audio cues were measured. Qualitative tests: The questionnaire is organized in two sets of Likert-scale items (derived from the System Usability Scale, and from IBM Computer Usability Satisfaction Questionnaire) and an additional set of open questions. |
| Najjar AB, Al-Issa AR, Hosny M. [34] | Dynamic indoor path planning for the visually impaired | 2022 | SAUDI ARABIA | To facilitate the navigation process for the VI, through developing a useful indoor navigation mobile application that can safely lead them to the desired destination. | 17 participants in total, 10 non-VI volunteers, and 7 VI participants. | No | users are involved in usability | No | Yes | No | not mentioned | The System Usability Scale has been used here to assess the comfort and acceptability of system use. |
| Connors E, Chrastil E, Sánchez J, Merabet L. [35] | Action video game play and transfer of navigation and spatial cognition skills in adolescents who are blind | 2014 | USA | we have developed a novel approach to train navigation and spatial cognition skills in adolescents who are blind. | Seven early blind adolescents aged between 16 and 17 years (3 males) familiar with the use of a computer key board interface | Yes | Yes | Yes | not described | not described | not described | Navigation success was assessed by the number of correct paths executed (expressed as mean percentage correct). Then, statistical tools were used for the analysis. Conversations, interviews, and usability evaluations, to gather input from potential end-users as part of the user-centred design employed, are mentioned but not described in the paper. |
| Al-Razgan M, Almoaiqeel S, Alrajhi N, Alhumeigani A, Alshehri A, Alnefaie B. [36] | A systematic literature review on the usability of mobile applications for visually impaired users | 2021 | Saudi Arabia | The goal of this paper is to provide an overview of the developments on usability of mobile applications for people with visual impairments based on recent advances in research and application development. | Regarding the reviewed articles: 1. Most studies faced difficulties regarding the sample size and the fact that many of the participants were not actually blind or visually impaired but only blindfolded; 2. A commonly discussed future work in the chosen literature is to increase the sample sizes of people with visual impairment and focus on various ages and geographical areas to generalize the studies. | N/A: SR |  |  |  |  |  | Reported: The most prevalent methods to evaluate the usability of applications were surveys and interviews. Focus groups were also used extensively in the literature. |
| Aziz N, Motalib AA, Sarif SM. [37] | User Experience of Interactive Assistive Courseware for Low Vision Learners (AC4LV): Initial Round | 2017 | Malaysia, | to investigate user experience of AC4LV in terms of information accessibility, navigationability, and pleasurable. | eight subjects with the average age 9 to 12 were involved. | No | Users are involved since the usability test of the prototype | No | The intention is declared by the authors. | Yes | Not described | Subjective feedbacks were obtained through observation and interview. Think-aloud Protocol. |
| Bateman A, Zhao OK, Bajcsy AV, Jennings MC, Toth BN, Cohen AJ, et al. [38] | A user-centered design and analysis of an electrostatic haptic touchscreen system for students with visual impairments | 2018 | USA | To detail the user-centered design and analysis of an electrostatic touchscreen system for displaying graph-based visual information to individuals who are visually impaired. Feedback AND to present the usability study of the AD developed | For the UCD process the participants included technology experts with visual impairments, principals and teachers of a school for VIPs. For the usability study, 12 VIP | Yes | Yes | Yes | Yes | Yes | The design team is not described. | Interviews, preliminary tests, Usability test, Video analysis |
| Colley M, Walch M, Gugenheimer J, Askari A, Ruzkio E. [39] | Towards Inclusive External Communication of Autonomous Vehicles for Pedestrians with Vision Impairments | 2020 | Germany | This work presents an inclusive user-centered design for VPC, beneficial for both vision impaired and seeing pedestrians. | N=6 participants (SD=4.44; range: 45 to 56) years old, researchers conducted a between-subject study with N=8 VIP and N=25 seeing people. | Yes | Yes | Yes | Iteration is described in the theoretical framework, not in the development process | Yes | Not mentioned | The workshop consisted of three phases: introduction, evaluation and an open discussion. Objective dependent variables: During each crossing, the system logged position, the lateral angle of the view, and the current duration of the crossing in relation to when the auditory crossing signal was given. Time on street. Subjective dependent variables: • affective state: on a 7-point semantic scale using the self-assessment manikin (SAM), • cognitive load: raw NASA-TLX [37] on a 20-point scale, • subscale Predictability and Trust in Automation of the Trust in Automation questionnaire by Körber |
| Fidyka A, Matamala A. [40] | Audio description in 360° videos Results from focus groups in Barcelona and Kraków | 2018 | Spain | To gather user feedback, through a series of focus groups, on how AudioDescription (and secondarily AST) could be integrated in immersive content, both from the perspective of producers and consumers. | Barcelona: 6 participants: 2 end users (partially sighted), 3 audio describers and 1 technical expert. Kraków: 6 participants (3 end users (blind) and 3 audio describers). Sex, age and other user's characteristics are reported. | Yes | Yes | Yes | Not declared | Yes | Not described | Focus groups |
| Shi L, Lawson H, Zhang Z, Azenkot S. [41] | Designing interactive 3D printed models with teachers of the visually impaired | 2019 | United States | In this paper, we present two studies that investigate how to design I3Ms as effective teaching aids. In both studies | 1st Workshop: 16 TVIs participated (1 with low vision, 2 blinds, 13 sighted). 2nd workshop: 19 TVIs (1 with low vision, 1 blind, 17 sighted). Age, experience and other participant's data are reported. | Yes | Yes | Yes | Yes | Yes | One of the researchers is an expert in education for students with VI. An accessibility specialist was included in the researchers team to assess the usability of the application before delivering it to the TVIs. | Workshops, brainstorm, meetings. The remote meetings between the researcher and each TVI, and the TVIs feedback to their students' were recorded. The data provided by the TVIs was the only one used because of "feasibility and privacy concerns". |
| Dough IA, Pontelli E. [42] | Non-visual navigation of spreadsheets | 2013 | USA | To measure the accuracy and the time needed when the user completes chart recognition tasks for bar, scatter, and line charts using the haptic/sound interface. The suitability of the feeling of the force feedback and the accuracy and effectiveness of the audio cues were also measured. Another goal was to compare the accuracy and time needed to navigate charts using the proposed system and using charts printed on tactile paper. | 7 unpaid students (3 females and 4 males) from New Mexico School for the Blind and Visually Impaired, 5 blind, 2 VI. | No | Involved in the usability test | No | Yes | no | Not described | 1. Quantitative measurements. 2. A questionnaire about each performed task and a questionnaire of the usability and accessibility of the system and potential suggestions to improve the system |
| Adebiyi A, Sorrentino P, Bohlool S, Zhang C, Arditti M, Goodrich G, et al. [43] | Assessment of feedback modalities for wearable visual aids in blind mobility | 2017 | USA | The purpose of this paper is to report on a study comparing two types of ETA outputs (speech or tactile) in a group of blind test subjects. | All subjects were blind with regards to functional vision. Subject code, age, gender and visual diagnosis are shown. For 1st Experiment : 11 participants, 2nd experiment: 10 participants | No | Users are involved in tests to compare audio and vibrotactile ETA outputs. | No | Intention declared | No | not described | A "person-in-loop" testing sessions. The variables measured were: Compliance (indoor and outdoor); Average Reaction Time, preferred walking speed with the cane (Control) and with each MFS (mobility feedback system). After all testing, SUS score. |
| Giraud S, Thérouanne P, Steiner DD. [44] | Web accessibility: Filtering redundant and irrelevant information improves website usability for blind users | 2018 | France | the main goal of the three experiments presented below was to test if this filtering provides a benefit in terms of cognitive load and usability according to the three usability criteria: effectiveness, efficiency and satisfaction | Participants were contacted via Internet through e-mail and forums dealing with disability. Fifty screen reader users voluntarily participated in these experiments. | No | Involved in the usability test | Yes | Yes | No | Not described | The NASA-RTLX questionnaire measured cognitive load, three usability criteria were assessed (effectiveness, efficiency, and satisfaction), questionnaire "System Usability Scale". Conducting semi-structured interviews would be necessary in order to collect the perceptions of users with blindness of such a tool (advantages, risks, opportunities) |
| Lopes SI, Vieira JM, Lopes OF, Rosa PR, Dias NA. [45] | MobiFree: A Set of Electronic Mobility Aids for the Blind | 2012 | Portugal | To design a set of complementary electronic mobility aids for the blind, to cover as much as possible, his personal, near and far spaces: an improved long cane; the concept of a pair of sunglasses focus in the detection of head-level obstacles and a directional speaker to obtain echo information of surrounding elements | Two blind people were invited | NO | In the paper only is stated user participation in the evaluation | No | Yes | No | Collaboration with the Department of Communication and Arts of the Aveiro University. In the design process, is reported. | In order to give some feedback about the device and improvement tips, 1 person used the cane for more than two hours, and the other used the cane during a week. Both gave positive feedback and some functional tips. |
| Mascetti S, Ahmetovic D, Gerino A, Bemareggi C, Busso M, Rizzi A. [46] | Robust traffic lights detection on mobile devices for pedestrians with visual impairment | 2015 | Italy | this paper focuses on the problem of recognizing traffic lights with the aim of supporting a user with VIB in safely crossing a road. | The experiment involved 2 blind subjects and 2 low-visioned subjects (unable to see the traffic lights involved in the experiment). | No | Users are involved in usability tests | No | Yes | No | The design team is not described. | A supervisor recorded whether the task was successfully completed and took note of any problem or delay in the process. The subjects were asked to answer a questionnaire. |
| Kammoun S, Parsehian G, Gutierrez O, Brihault A, Serpa A, Raynal M, et al. [47] | Navigation and space perception assistance for the visually impaired: The NAVIG project | 2012 | France | The aim was to design and evaluate a powerful assistive device combining both micro- (sensing the immediate environment) and macronavigation (reaching a remote destination) functions. | in collaboration with the Institute of Young Blinds (CESDV-LIA, Toulouse). A panel of 21 VI users has been involved during all design steps. These users have been selected using several criteria, including motivation to participate in the project, self-sufficiency in O&M, and some degree of practice with new technologies. The subjects in the panel were between 16 and 65 years old; eight females and 13 males | Yes | Yes | Yes | Yes | Yes | NAVIG brings together two research laboratories in computer science and information technology and one research laboratory in human perception. IRIT-Elipse, the project leader, is an interdisciplinary research group in Human Computer Interaction (HCI). | The brainstorming sessions and discussions with VI users highlighted that an ideal system has to provide the best-suited level of audio guidance information |
| Lee K, Hong J, Jarjue E, Mensah EE, Kacori H. [48] | From the Lab to People's Home: Lessons from Accessing Blind Participants' Interactions via Smart Glasses in Remote Studies | 2022 | United States | To explore ways to overcome challenges associated with remote observations of blind participants' interactions via video conferencing with smart glasses | 12 blind participants, serving as a case study. | Yes | Yes | Yes | Yes | No | Our team, including four sighted researchers and one blind researcher. | videos |
| Younis O, Al-Nuaimy W, Rowe F, Alomari M. [49] | A Smart Context-Aware Hazard Attention System to Help People with Peripheral Vision Loss | 2019 | UK | This paper presents a new context-aware hybrid (indoor/outdoor) hazard classification assistive technology to help people with peripheral vision loss in their navigation using computer-enabled smart glasses equipped with a wide-angle camera. | 5 visually impaired | Yes | Yes | Yes | Intention declared | Yes | Authors declared research collaboration with the Department of Health Services Research | 3 Questionnaires: 1 about the challenges VIP face that would affect their QoL and their independent navigation; 1 about the hazardous situations while navigating. Dataset for hazard detection and classification; 1 Feedback regarding the system concept and alerts. Group meeting with patients. |
| Feiz S, Borodin A, Bi X, Ramakrishnan IV. [50] | Towards Enabling Blind People to Fill Out Paper Forms with a Wearable Smartphone Assistant | 2021 | USA | To present PaperPal, a wearable smartphone assistant which blind people can use to fill out paper forms independently. | For the WOZ pilot study: 8 blind participants. For the user study: 8 blind participants. Gender, age, skills, and other participants' data were reported. | Yes | Yes | Yes | Yes | Yes | not described | semi-structured interview to gather demographic data, reading/writing habits, and prior experiences with assistive smartphone apps. The experimenter making notes throughout the video recorded session. A single ease question to each participant to rate the difficulty of assembling the holder and completing each form, on a scale of 1 to 7 open-ended discussion |
| Real S, Araujo A. [51] | Navigation Systems for the Blind and Visually Impaired: PastWork, Challenges, and Open Problems | 2019 | Spain | To re-evaluate the perspective of navigation systems for the BVI in this new context, attempting to integrate key elements of what is frequently a disaggregated multidisciplinary background. | N/A | N/A: Literature review |  |  |  |  | not described | N/A |

### The design is based upon an explicit understanding of users, tasks, and environments

In reviewing the application of this principle in the available evidence, we sought compliance with the following points: identification of user and stakeholder groups, understanding of users’ needs and description of the context of use: “specified users, having specified goals, performing specified tasks”.

All the reviewed articles reported the participation of VI users, even though six studies complemented their samples with non-VI participants. Regarding sample sizes, the number of participants varies from two to 128 (55 VI, 71 non-VI), being 12 participants the number that appears the most often. Only seven articles reported sample sizes with more than 12 users. There is no clear rationale for why and how these samples were designed and selected, especially when considering the quantitative methodologies. Only one paper, Nimmolrat et al. reported the sampling technique and inclusion criteria for a sample of 60 participants [27].

Quantitative standards regarding sampling were not observed either. Mascetti et al, reported difficulties in recruiting test subjects with VI or blindness [33]. Under that argument, the paper added non-VI participants to the study and reported on results that merge data from both non-VI individuals and VI individuals. Najjar et al., whose sample consisted of 10 non-VI and seven VI participants, noted their limitations without being specific or addressing bias on the data analysis [34]. Connors et al. acknowledged that their sample size (7 blind adolescents) was “relatively small” and limited to carry out a correlation-based analysis [35].

Absence of persons with VI tend to be common, Al-Razgan et-al., reports that “most studies faced difficulties regarding the sample size and the fact that many of the participants were not actually blind or visually impaired but only blindfolded” [36].

When qualitative methods were applied, no standard sampling techniques nor quality assurance practices for qualitative sampling were reported, e.g. characterization of patterns, and variations among the participants, data saturation. Studies such as Aziz et al., argued that the sample size (8) was sufficient considering its qualitative nature, without providing any further rationale [37]. Furthermore, Conradie et al. claimed that two focus groups (sample sizes 9 and 12) “served to reveal the experiences and knowledge of blind persons” to the researchers which make it possible to sketch broad user needs within the target group and specifying varying degrees of mobility needs and assistive device demands [21].

In terms of characterising users, participants’ details were poorly described. The sex of the sample members was reported by 19 papers, the same number of articles stated the age of the participants, four of the studies were addressed to minors (8-17); two other papers reported participation of teenagers and young adults; the rest of the studies included adults only in different age ranges (18-78). Other types of data reported were: the participant’s skills related to the use of the designed AT (14 papers); the education level of the participants (10 papers), their occupation (five studies), the number of years lived with visual disability or the year when the disability was acquired (five papers), and the use of aids (two studies). Environment of use was often not mentioned. Only one paper reported an analysis of the physical environment in which the product will be used, the user’s social and organisational milieu and the technical environment and associated technical constraints [24].

As for stakeholder identification, 8 papers mentioned the involvement of stakeholders. Four papers declared the participation of academics (principals, teachers and O&M trainers), three studies included representatives from disabled people’s organisations, and four studies included technology experts [23-26, 38-41]. Bateman et al., included all the above [38].

### Users are involved throughout the design and development

All of the reviewed articles reported user involvement, but rarely throughout all of the stages of the design process. Four of the studies present data from the design phases in which participants were actively involved [21, 25, 39–40]. Their involvement included “in-depth requirements analysis” through users and stakeholders’ feedback through a series of UCD comprehensive methods. Mattheiss, et al. first centred on analysing the requirements in the areas of Orientation and Mobility (O&M) training and accessible video game play to later work on the first iterations of the design, implementation, and evaluation of the developed game editor [25]. In this case, authors declared the involvement of children (end-users) as design partners.

User participation in the final stages, namely for evaluating the solutions, was stated by 5 studies: 4 in usability testing [37, 42–44] and 1 in field testing [45]. Involvement, both in the design and testing phases, was reported by 14 studies [23, 26–28, 33–35, 38, 41, 46–50]. Although, it is pertinent to point out that: on the one hand, some studies mention the involvement of the participants at the beginning and at the end of the process, but not in all the stages of the process. On the other hand, how users were involved tend to be unclear and reporting of such involvement tends to be rather superficial, for example, Najjar et al. mentioned the identification of potential users’ requirements, but these are not presented in the article [34], instead a previous study is referred to. Nimmolrat et al. provide a better description of users’ participation during the design process [27]. Ntakolia et al. [24] detailed user’ś participation in the design and development phases, however, usability testing was done with blindfolded non-VI participants only [24].

### The design is driven and refined by user-centred evaluation

The use of user-centred evaluation tends to be more explicit, explained and applied in studies that used qualitative methods, such as behavioural observation, think aloud techniques, in-depth interviews, and focus groups, among others these kinds of evaluation methods allow the user’s perspective to be addressed early [21, 23–27, 37–38, 40, 50]. The analysis of the context of use could determine the user’s needs against which the preliminary design solutions will be tested.

Usability evaluations reported on the evidence collected included both quantitative and qualitative methods. There is a stronger emphasis on quantitative scales to assess usability, such as the System Usability Scale. In addition, usability was assessed in terms of the performance of the technology and other quantitative variables related to efficiency (time) and effectiveness. Except for the studies by Najjar et al. and Giraud et.al, the studies which applied quantitative methods for usability testing, also reported userś feedback without specifying the methods used to gather that data [34, 44], e.g. Lopes et al., stated that subjects had the chance to use the device and were asked to give feedback, but did not describe the methods for data collection [45].

There is a lack of real-world scenarios when evaluating AT. Some studies claim that this was because they are focused on preliminary solutions, and in some others, because the study has pure research and non-commercial orientation [25, 28, 34, 44]. Ntakolia et al., excluded users from its evaluation process of the prototype, reporting that future research would include VI users for the usability test [24].

### The process is iterative

In reviewing compliance with iteration, which dictates the iterative repetition of a sequence of steps until the desired outcome is achieved, it is important to remember that not all the included articles report the complete UCD process, but some focus on design and several, as presented initially, are limited to the usability evaluation of prototypes. Thus, all the studies reported iterations, or the intention to make them, based on the feedback gathered from studies’ participants. Iteration involves not only the prototype but also the descriptions and specifications, the refinement of information from the feedback obtained during the development process and in usability testing, is also considered. From this perspective, a noteworthy study on this subject is that of Bateman et al. where design and re-designs were submitted to preliminary tests with expert users [38],. Finally, the usability test conducted with 12 students confirmed that the previously expressed needs regarding accessibility and effectiveness were met. The authors went beyond mentioning that an iterative UCD process was carried out, in fact, they went on to explain the information gathered and the stakeholder’s characteristics through every round of interviews. The iterations made and the preliminary test results were also detailed. In other words, iterations were placed in the context of use.

Another interesting example is the study by Shi et al., where two studies were conducted to understand how to design effective, interactive 3D models for education purposes for blind students [41]. In the first study, two design workshops were performed with teachers of VI students (TVIs) in which suggestions from conceptual designs were aggregated. Then, the second study was performed with three teachers of VI students, not only to design, but to deploy sample interactive 3D models over seven weeks. In-depth work with individual TVIs, and deployment of interactive 3D models in their classrooms were reported by the researchers, resulting in improvements to the prior system and mobile application development that supports the use of interactive 3D printed models in an educational setting. Additionally, the authors stated that based on the feedback from the second study, the mobile application could be further improved.

Although in less depth than the cases previously discussed, Conradie et. al and Mattheiss et al. highlighted the importance of rapid prototyping in the execution of iterations [21, 25]. Adebiyi et al. and Feiz et al. emphasised the effectiveness of the “Wizard of Oz” technique in achieving development improvements [43, 48]. This technique consists of a tactic used for low fidelity prototyping in which the participant receives instructions in order to perform tasks while testing a prototype, and a human simulates the behaviour of the completed AT. For example, for a navigation device, a person will simulate the task that the device will perform by providing vocal instructions to the users.

In turn, Doush and Pontelli reported “iterative modifications have been applied to the system based on empirical studies carried out with the participation of sighted and blind users”. However, they do not describe the iterations performed or how these studies were conducted, nor do they explain why non-visually impaired participants were involved [42]. Likewise, Mascetti et. al, stated that “during the design of the auditory guiding modes several test subjects were asked to use the application and provide feedback” this was done via informal test [33].

### The design addresses the whole user experience

ISO 9241 stresses that usability goes beyond “making products easy to use”, by considering perceptual and emotional aspects as keys to understanding the user’s experience from their own perspective.

Still, several studies assessed usability mainly by considering parameters such as ease of use, accessibility, or satisfaction with the device [33-34, 42-44, 46]. These studies applied quantitative scales. To have information to improve the device, three papers reported to have included questionnaires or open-ended questions (not described in the papers) [33, 42, 46]. In yet another case, in which only System Usability Scale (SUS) was applied, feedback from users was reported as results of “anecdotal comments” [43].

Mascetti et al., reported as a result of feedback from participants after evaluation of the prototype, that they did not desire to hold a mobile phone in one hand while holding a cane in the other [46]. This type of information evidences that the characterisation of users’ needs and preferences was not carried out at an early stage and therefore, users’ previous experiences and perspective were not addressed.

Other feedback refers to the time the user needs to get familiar with the device, the need for more training time was expressed by the participants in the studies conducted by Doush and Pontelli and by Mascetti et al. [42, 46]. It was also stated by Najjar et al. [34]. On the other hand, although Giraud et al., did not include feedback within the methods or results, they did express the future need of conducting semi-structured interviews “in order to collect the perceptions of users with blindness of such a tool (advantages, risks, opportunities)” [44].

Alternatively, preferences and expectations were mainly assessed in the studies of Mattheiss et al., and Aziz et al.: design for skills development in VI children through an interactive learning material and a videogame, respectively [25, 37].

Furthermore, Sánchez obtained feedback from users regarding their emotions [26]. Colley et al., also considered affective state variables, namely ‘control over the situation’ in the analysis [39]. Nimmolrat et al., assessed satisfaction with the functionality of the application through interviews [27]. Finally, eight studies (28.57%) based their development on the available literature and did not include collecting any empirical data.

### The design team includes multidisciplinary skills and perspectives

The large majority of studies did not report multidisciplinary skills and perspectives. Only two studies described the research team. Facanha et al., stated that the design team included four undergraduate students in the computer sciences, an assistive technology researcher and an ophthalmologist [23]. Shi et al., mentioned that one of the researchers of the team is an expert in education for students with visual impairments, and that they included an accessibility specialist [41]. Nevertheless, some authors did report collaborations: Kammoun et al., declared the participation of different engineering research groups in human perception, human-computer interaction, audio and acoustic, and spatial cognition and perception. Further, the authors mentioned the project leader, is an interdisciplinary research group in Human Computer Interaction [47]. Other reports of collaborations outside the engineering team are: Lopes et. al, who mentioned a collaboration with the Department of Communication and Arts of Aveiro University [45] and Younis et al., who declared a research collaboration with the Department of Health Services Research in the UK [49].

## Discussion

The literature reports a growing trend in the application of user-centred design in the development of assistive technology for the visually impaired persons [51]. However, the results show that evidence on the effective implementation of UCD with VI on the design of AT is scarce. Publications show that the principles of the ISO 9241-210 (user-centred design) tended to be not fully applied, despite being called guiding principles and despite the increasing availability of models and frameworks that could facilitate their application [52].

### The focus was on the system requirements, not its user

The information on the system architecture reported in the state of the art in the analysed studies, was prioritised over the participants’ needs with respect to AT. For these articles, it was common not to find specifics on sample size calculation and participant’s selection. Participation of potential users was low and was accompanied by a superficial description of their profiles. Users’ involvement was reported mainly in usability assessment at the end of the process rather than in design phases, and when users’ involvement was reported in design phases, it was usually not thoroughly described. This contrasts with the extent in which technological aspects of the development were informed. Also, a stronger focus on the verification of the system, over its validation, was observed. According to quality management standards, such as the ISO 9001, independent validation and verification (V&V) processes need to be performed to determine if a developed system meets the defined requirements and specifications and fulfils its intended purpose [53–54]. Specifically, the verification process focuses on the system’s requirements (“Did we developed the system right?”) while the validation process focuses on the system’s worthiness, i.e., if it fulfils its intended purpose, user expectations, etc. (“Did we developed the right system?”) [55].

For usability assessment, most studies used surveys and quantitative methods to gather information. Though standard parameters of quality on those methods, such as rationale for power and limitation of the sample size calculation, were not met. Feedback from users, when present, took the form of “informal”, “casual” or “anecdotal” data. Moreover, in these studies iterations are often mentioned in the evaluation phase and not in the design phase. At this point, it is important to emphasise that according to the ISO 9241-210, iterations should be done throughout the process and not only at the evaluation stage.

Regarding usability, ISO 9241-210 states the need to go beyond the concepts of ease of use and effectiveness, and to incorporate userś experience. In this perspective, the standard recommends to consider the userś skills, habits and personal goals, as well as emotional aspects and experiences of previous solutions or alternatives. Notwithstanding, the studies under consideration fall short in assessing the whole users’ experience, there was little or no information on the social and environmental context in which these devices were intended to be used. In this regard infrastructural constraints such as internet availability, road safety or social aspects like stigma are not accounted for.

### Moving towards better understanding of the final users

Characterization of user needs was often unstructured, lacked robustness or tended to be underreported. This trend has been previously observed in the study of requirements elicitation techniques [52]. This was also observed in the present review. Among the reasons given to justify such behaviour are limited resources, time and endeavour to conduct a thorough requirements assessment process [52]. Similarly, some of the studies in this review reported major logistical challenges in recruiting participants.

There is a growing number of articles that seek to better engage with users of AT. This was generally achieved either because they took care to obtain larger samples under previously defined selection criteria or because they selected more appropriate methods (qualitative or mixed) with respect to the objective pursued, or due to both reasons [24-25, 27, 47]. It is also pertinent to highlight the importance of having included stakeholders in these studies [23-25, 27, 40-41, 47-48].

Researches did not fully apply all ISO 9241-210’s principles. However, it can be argued that a better compliance to the first principle (*the design is based upon an explicit understanding of users, tasks, and environments*) increased the probability of applying the subsequent four principles. The fact that some studies integrated participatory design approaches into the methodology boosted the involvement of participants in the whole process [23, 25, 27–28, 41, 48]. The participation of both potential users and stakeholders in the early design phases and throughout the process, as well as the type of instruments applied to collect information, allowed the design to be “driven and refined by user-centred evaluation”. Iterations were reported both in the information collected to guide the design, and in the prototypes.

Regarding usability, in addition to the application of validated surveys and the analysis of system performance parameters, qualitative methods were used to obtain feedback from users in a more systematic and deeper way, and to compare it with the initial information from the context of use. In some cases, the user experience was assessed in a more comprehensive way by considering the emotions evoked through interaction during the prototypes assessment [25, 37, 39].

### Multidisciplinary skills and perspectives

So far, the application of five of the six principles of the ISO 9241-210 in the reviewed articles has been discussed. Regarding the application of the last principle “The design team includes multidisciplinary skills and perspectives” in the reviewed studies, this is where the least evidence was reported. Although the standard does not define the need for broad heterogeneity of the disciplines involved in the process, since it is designed to guide processes of different natures, it is understood that disciplines from diverse fields are needed to elicit and comprehend userś needs and to address the system’s requirements. Multidisciplinary teams would not only allow dealing with the issues related to technology but also those that have to do with the users’ functionality, and above all, it would facilitate the mixed methodological approach.

As for the literature, in addition to designers and engineers, it is proposed in specific cases to work with clinicians, health professionals or rehabilitation professionals and with Commercial specialists [51–53].

### From user centred to person centred

Literature addresses two streams of user-centred design, one in which the “user” is placed at the centre of the design process and another, which focuses on the “person” [56]. The main difference lies in the fact that the first considers the interaction between the user and the product and “is concerned with ensuring that artifacts function as intended by the designers”. While the latter also accounts for context-determined interactions and focuses on “enabling many individual or cultural conceptions to unfold into uninterrupted interfaces with technology.” Giacomin et al., add that products acquire meaning when used by persons, and that it is the understanding of that meaning that should guide the design. In their words, “the natural focus of questions, insights and activities is on the people for whom the product, system or service is intended, rather than on the designer’s personal creative process or on the material and technological substrates of the artefact” [56]. We might say that most articles followed the first trend (user-centred), while there is less evidence of the second understanding (person-centred) when it comes to AT.

## Conclusion

This review explores how well the principles of ISO 9241-210 are applied in the case of AT. As for the implications, on the one hand, it highlights that the application of the UCD approach is not standardised in the field of AT design for the visually impaired. Although there is a standard that guides the implementation of the approach and has been thoroughly reviewed by ISO experts, it has not been embraced in this field. On the other hand, there is also a lack of methodological rigour in understanding the needs of users in their context, revealing that people are not at the centre of the process in a generalised manner. Furthermore, it is evident that the developments are carried out in a disarticulated manner, so that recommendations made by international authorities on the subject, such as those given by WHO in GREAT [7], are also disregarded.

Based on these findings, we emphasise the need to pay greater attention to the principle: ‘Users are involved throughout the design and development’, meaningfully engaging with users would lead to better identification of their needs and preferences. It shall also improve the possibility to have better recruitment procedures, representative samples and more representative and robust results.

Engaging with users will require a broad level of expertise and full implementation of ISO 9241-210 principle 5.7 ‘The design team includes multidisciplinary skills and perspectives’. Transdisciplinarity could have reduced methodological flaws observed in the literature today. Transdisciplinarity shall open the possibility of cross-fertilization between the different fields of knowledge and in conjunction with people with visual impairments as potential direct users and with their stakeholders. The application of this principle shall enable design teams to include not only diverse classes of engineers, but also designers, health and rehabilitation professionals, social scientists including disability scholars, anthropologists, and economists, among others. Such teams shall be better equipped to develop and apply a range of methodologies that understand the social, physiological, cultural and technological needs of the target users and develop AT that responds to them. The strengthening of this last principle of the standard would lead the work towards the consolidation of adequate methodologies to gain a better understanding of how AT could enable visually impaired people to live the lives they would like to live.

Design of AT should be focusing on enhancing the user’s agency, bodily integrity, and capabilities, and not trying to “fix disabled bodies”. Evidence collected suggests that assistive technology has focused on functional deficiency solely, namely impairment rather than in enhancing wellbeing for its users [6, 57]. The latter seems to be the prevalent approach today as users are for the most part not meaningfully included in the design and development and only call to test a final product that aims to provide a ‘solution’. “Nothing About Us Without Us” should resonate with the design, development and implementation of any technological development that concerns persons with disabilities.

## Data Availability

All data produced in the present work are contained in the manuscript

## Supporting information

S1. PROSPERO PROTOCOL

S2. PRISMA 2020 Checklist

S3. Quality assessment for the studies

